# Regional endemicity of toxigenic *Vibrio parahaemolyticus* lineages associated with foodborne illness in Australia

**DOI:** 10.64898/2026.08.19.26360778

**Authors:** Jake A Lacey, Claire E Hedges, Anne E Watt, Valeria A Torok, Cheryl Jenkins, Neil Franklin, Daniel R Knight, Emily Fearnley, Karolina Mercoulia, Lito E Papanicolas, Rikki M A Graham, Lex Ex Leong, Amy V Jennison, Vitali Sintchenko, Benjamin P Howden, Norelle L Sherry, Alison Turnbull

## Abstract

Gastrointestinal *Vibrio parahaemolyticus* infections, primarily associated with consumption of oysters, are emerging in Australia, where previously little was known about the disease and epidemiology. Following a multijurisdictional outbreak in 2021 and additional smaller outbreaks in subsequent years, an opportunistic whole genome sequencing study was undertaken to characterise human illness-causing strains in Australia. Through a multijurisdictional collaboration that bridged research, government, pathology service providers, aquaculture and clinicians, 676 *V. parahaemolyticus* genomes were contributed for analysis from human clinical, food, and environmental samples. We identified ST36, ST50 and ST417 as the dominant multi-locus sequence types causing gastrointestinal illness nationally. Phylogeographic contextualisation of Australian *V. parahaemolyticus* sequences within the global dataset indicates the Australian and New Zealand ST36 strain originated from a single point of introduction from the US Pacific-Northwest and is now circulating locally. In contrast, ST50 and ST417 appear to be endemic across Australia, with multiple lineages co-circulating. These findings establish a baseline for future outbreak investigations of *V. parahaemolyticus* in Australia and the consolidation of Australian data provides a critical platform for ongoing research, public health surveillance and risk mitigation.

## Introduction

*Vibrio parahaemolyticus* is a Gram-negative, halophilic bacterium ubiquitous in marine and estuarine waters worldwide^1^. Human infection most commonly arises from ingestion of raw or undercooked seafood, particularly filter-feeding bivalve shellfish, such as oysters, and manifests as gastroenteritis, although wound infections and, in severe cases, invasive disease^2,3,4,5^ may also occur. Pathogenic potential is largely determined by two hemolysin genes, thermostable direct hemolysin (*tdh*) and thermostable-related hemolysin (*trh*), which together define a small pathogenic subpopulation within the broader population^1^. Environmental proliferation may occur when water temperatures are above 14–19°C, and rising ocean temperatures under climate change are anticipated to expand both the geographic range and seasonal window for *V. parahaemolyticus* occurrence in shellfish growing areas^2–5^.

*V. parahaemolyticus* is a leading bacterial cause of seafood-associated gastroenteritis worldwide, causing hundreds of thousands of infections annually and exhibiting increasing incidence in temperate regions due to climate-driven expansion of suitable marine habitats. Recent surveillance data recorded 499 laboratory-confirmed cases in the United States during 2024^6^, more than 23,000 confirmed infections in China over the past decade^7^, and a growing number of large outbreaks across Australia and Europe.

Australia recorded predominantly small *V. parahaemolyticus* outbreaks in humans, with four documented events affecting 24 people in total between 2002 and 2019^8^. This pattern likely reflects a low baseline incidence and/or under-reporting due to limited routine laboratory testing, and the pathogen’s variable notification status across Australian states and territories prior to 2025. The vehicle for these outbreaks (where known) was oysters. Oyster production predominantly occurs in temperate waters in New South Wales (NSW), South Australia (SA) and Tasmania (Tas) (USD$55M, $41M and $26M, respectively)^9^ with minor production in Western Australia (WA) and Queensland (Qld). Most of these oysters are sold domestically (99%), with trade frequently moving across state borders^9^.

Since 2021, a further three oyster-associated outbreaks have been documented, spanning multiple Australian states and territories and resulting in over 310 laboratory-confirmed illnesses^10^. Two of these events were attributed to the pandemic sequence type (ST)36, one in 2021 and one in 2024^11,12^. A polyclonal outbreak in 2021 was observed, with two sequence types, ST50 and ST417 identified as the causative agents^10^. Whole-genome sequencing (WGS) of clinical and oyster isolates confirmed that ST36 and ST50 strains carried both *tdh* and *trh*, whereas ST417 isolates harboured *trh* alone, underscoring the emergence and multisource dissemination of virulent sequence types within Australian shellfish production systems. Since 2019, New Zealand (NZ) reported its first of a series of domestic outbreaks attributed to *V. parahaemolyticus* ST36 and ST50 associated with consumption of raw or partially cooked mussels among a range of other seafoods^13^

With an increased rate of disease being observed in Australia, it has become clear there are critical knowledge gaps regarding the national distribution of *V. parahaemolyticus*, the environmental and ecological drivers precipitating outbreaks, and the molecular markers best suited as targets for development of rapid detection methods of analysis for high-risk strains. To bridge these gaps, and in response to the recent increase in reported outbreaks, a coordinated national collaboration drawing contributions from Australian states and territories has been established. To create a public database of sequencing data, the collaboration opportunistically collected and sequenced available *V. parahaemolyticus* isolates recovered from clinical cases, oysters, and other environmental/food samples. By integrating WGS data from Australian isolates with publicly available genomes internationally, we aim to contextualize the genomic diversity and transmission dynamics of outbreak-associated sequence types.

In this study, we focus on the genomic data from clinical, food and environmental isolates collected. We present a comprehensive genomic epidemiological analysis of the major *V. parahaemolyticus* sequence types (ST36, ST50, and ST417) linked to recent Australian outbreaks^10^, situating them within the broader national and global phylogeographic landscape^14–16^. We leverage high-resolution phylogenies, virulence gene profiling, and population structure assessments to trace introduction and dissemination pathways across Australian jurisdictions and compare Australian outbreak strains to international lineages of concern. These insights will inform targeted risk assessments, enhance early detection frameworks, and guide evidence-based interventions to safeguard public health in Australia’s burgeoning shellfish industry.

## Methods

### Setting and Study Design

This retrospective study aimed to characterise and analyse, through WGS, *V. parahaemolyticus* isolates collected by project collaborators from areas across Australia, which is divided into eight states and territories. To form the collaboration, health departments, diagnostic laboratories and environmental researchers in each jurisdiction were invited to contribute WGS data or identify isolates that could be sequenced and included in the dataset. High-level metadata was sought for each isolate being included in the study to provide context on isolate source (species, year of isolation and geographical location of sampling). The study protocol and metadata collection was designed to be generic and the data collected was exempt from human research ethics review through the University of Tasmania.

### Sources of Isolates

Sequences from clinical isolates collected between 2004 and 2025 were analysed, spanning the transition from jurisdiction-specific notification (Northern Territory (NT), TAS, WA, and SA from 2016; notification in other jurisdictions was not mandatory) to mandatory national notification implemented in 2025. Clinical isolates were predominantly associated with gastrointestinal disease from 2021 onwards, including the 2021 and 2024 outbreaks mentioned previously. Additional sporadic (non-outbreak) cases of gastroenteritis and wound infections were also included. Food and environmental isolates were also included, primarily from targeted oyster surveys, market surveillance, and routine food testing. Although efforts were made to obtain a local representation of relevant isolates, the opportunistic nature of data inclusion resulted in uneven representation and incomplete national coverage.

### Whole-Genome Sequencing and Quality Control

DNA extraction and WGS of study isolates was performed at various laboratories around Australia with sequencing undertaken on Illumina MiSeq100, NextSeq500 and NextSeq550 platforms. Quality control, sequence identification, genome assemblies, annotation, and assessment of genome metrics were conducted on the combined national dataset using the *bohra* microbial genomics pipeline (https://github.com/MDU-PHL/bohra) using *seqkit* v2.1.0, *KMC* v3.2.1, *Kraken* v2.1.2, *shovill* v1.1.0, *SPAdes* v3.13.2, P*rokka v*1.14.6.

### Genotyping and Antimicrobial Resistance Detection

*In silico* serotyping was performed using *Kaptive3*^17^ with *V. parahaemolyticus* O and K schemes^18^. *In silico* multi-locus sequence typing (MLST) using the “vparahaemolyticus” scheme^19^ was conducted to assign a sequence type (ST) using *mlst* v2.23.0 (https://github.com/tseemann/mlst). Species-level clustering was conducted with *popPUNK* v2.4.0^20^ (database was built from 10,437 publicly available genome assemblies). Antimicrobial resistance genes were detected from genome assemblies using *abriTAMR* v1.0.14^21^ with default parameters. Genes associated with virulence were detected using *abricate* 1.0.1 using the 2025 virulence factor database^22^ with an 80 minimum coverage and an 80 minimum identity threshold.

### Variant Calling and Phylogenetic Inference

Identification of core genome single-nucleotide polymorphisms (SNPs) against reference genomes (Supplementary Table) was performed using *snippy* v4.4.5 (https://github.com/tseemann/snippy, applying minfrac 10 and mincov 0.9). For WGS data where only genome assemblies were available, snippy contigs mode (–ctgs) was utilised. The SNP variants identified by *snippy* were used for phylogenetic reconstruction using maximum likelihood (ML) in *IQtree2* v2.1.425^23^ with constant sites corrected, 1000 bootstraps, and a generalised time-reversable model of evolution GTR+F+G4. *Gubbins* v2.4.1^24^ was used for recombination detection and bacteriophage detection was conducted using *PhastAF* (https://github.com/tseemann/phastaf) using the Phaster database^25^. Phage and recombinogenic regions were masked from the core alignments for which a recombination-free ML phylogeny was conducted using the same parameters in *IQtree2. Treetime* (v0.11.3)^26^ was utilised for ancestral state reconstruction and inference of timed tree phylogenies applying the --covariation --stochastic-resolve --coalescent skyline –confidence parameters, isolates determined to be outliers through these measurements were removed from the analysis. Phylogenies and skyline plots were processed and visualised in R v4.3.1 using the ggtree v3.8.2, treeio v1.24.3, ggplot v3.5.0, dplyr v2.4.0, readr v2.4.1 and ape v5.7-1 packages.

### International Context Isolates

To provide phylogeographic context for this dataset, we uploaded all read sets from this study to the NCBI Sequence Read Archive (SRA) and integrated those datasets into the Genome Detection Portal (GDP). We searched all isolates to determine their corresponding cluster designations. For each identified cluster, we downloaded additional publicly available raw sequencing reads where key metadata (date of isolation and country of origin) were also available. All public read sets were processed using the same quality control thresholds as mentioned previously and incorporated into the ST-specific analyses. Additionally, we downloaded 10,437 genome assemblies from Refseq and GDP to provide context to our study (Supplementary Table 1).

### Code and reproducibility

Whole genome sequencing reads for this project were stored in the following Bioprojects PRJNA1466780, PRJNA1129299, PRJNA1499355, PRJNA1131944, PRJNA856407, PRJNA1083213, PRJNA475810, PRJNA1194145, PRJNA1300975, PRJNA670297, PRJNA1514178 and PRJNA915130. Code for reproducibility purposes is stored in github at repository https://github.com/JA-Lacey/SafeFish-Vpara

## Results

### Clinical cases and environmental reservoirs of concerning *V. parahaemolyticus* lineages in Australia

This study analysed 676 *V. parahaemolyticus* genomes collected across Australia between 2004 and 2025, including clinical cases (n = 363), environmental/food sources (n = 299), and unknown sources (n = 14). Most isolates were collected during the outbreak years (2021–2024, n = 600) (**Figures 1 and 2**). Seven jurisdictions contributed data, and all but one provided both clinical and environmental/food isolates. We identified 83 unique MLST combinations, and a further 100 isolates with novel MLST profiles. Capsular typing revealed 17 O-serotypes and 75 K-serotypes. Clinical isolates were predominantly from gastroenteritis/faecal samples associated with outbreaks. Environmental/food/animal isolates were from targeted sampling of oysters (n = 249), with smaller numbers from other seafood (prawns, snails, mussels; n = 20), miscellaneous foods (n = 25), and water samples (n = 4).

**Figure 1.**
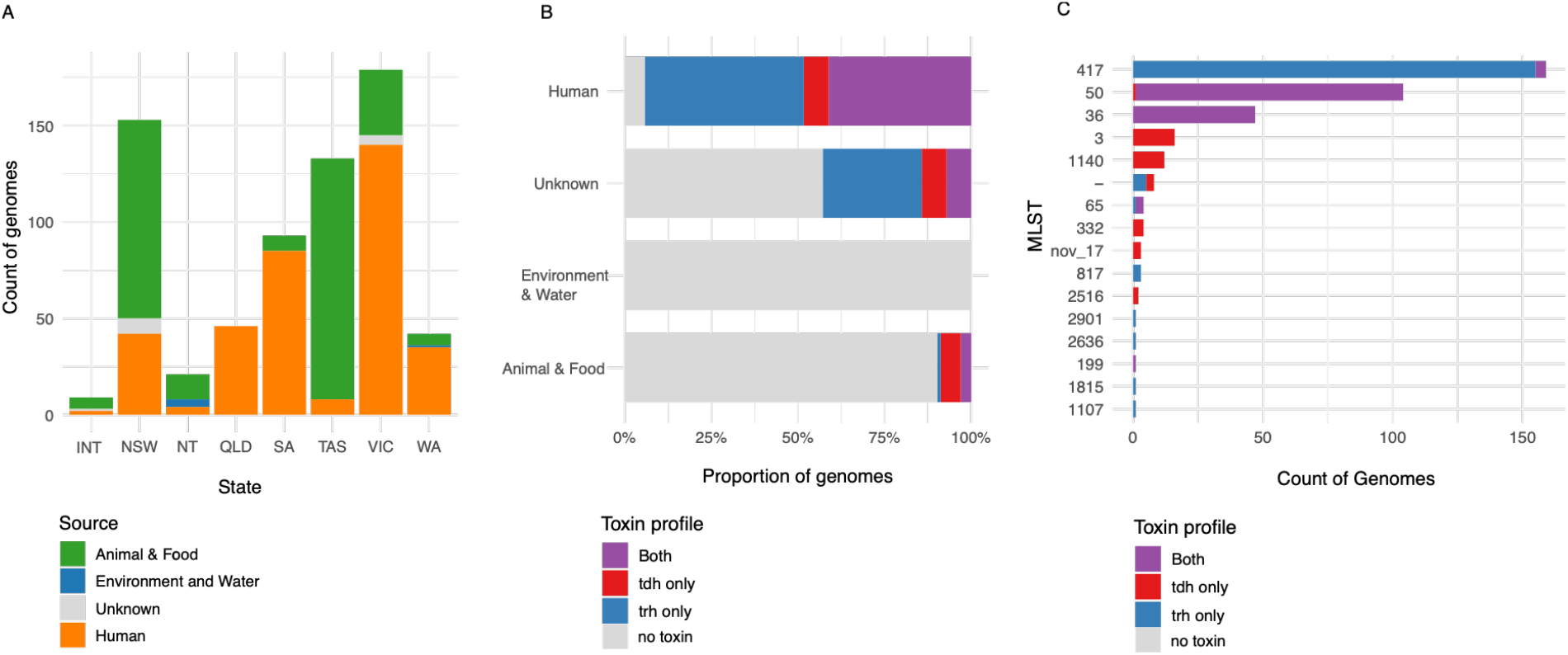
Characteristics of included isolates. **A**. Counts of V. parahaemolyticus isolates coloured by source (Animal and Food (green), Environment and Water (blue), Human (orange) and Unknown (grey)) from each contributing Australian state and territory, **B**. proportion of genomes by source type found to be toxin carrying (tdh, trh or both) and **C**. showing the counts of V. parahaemolyticus genomes attributed as toxin carrying strains by MLST.

**Figure 2.**
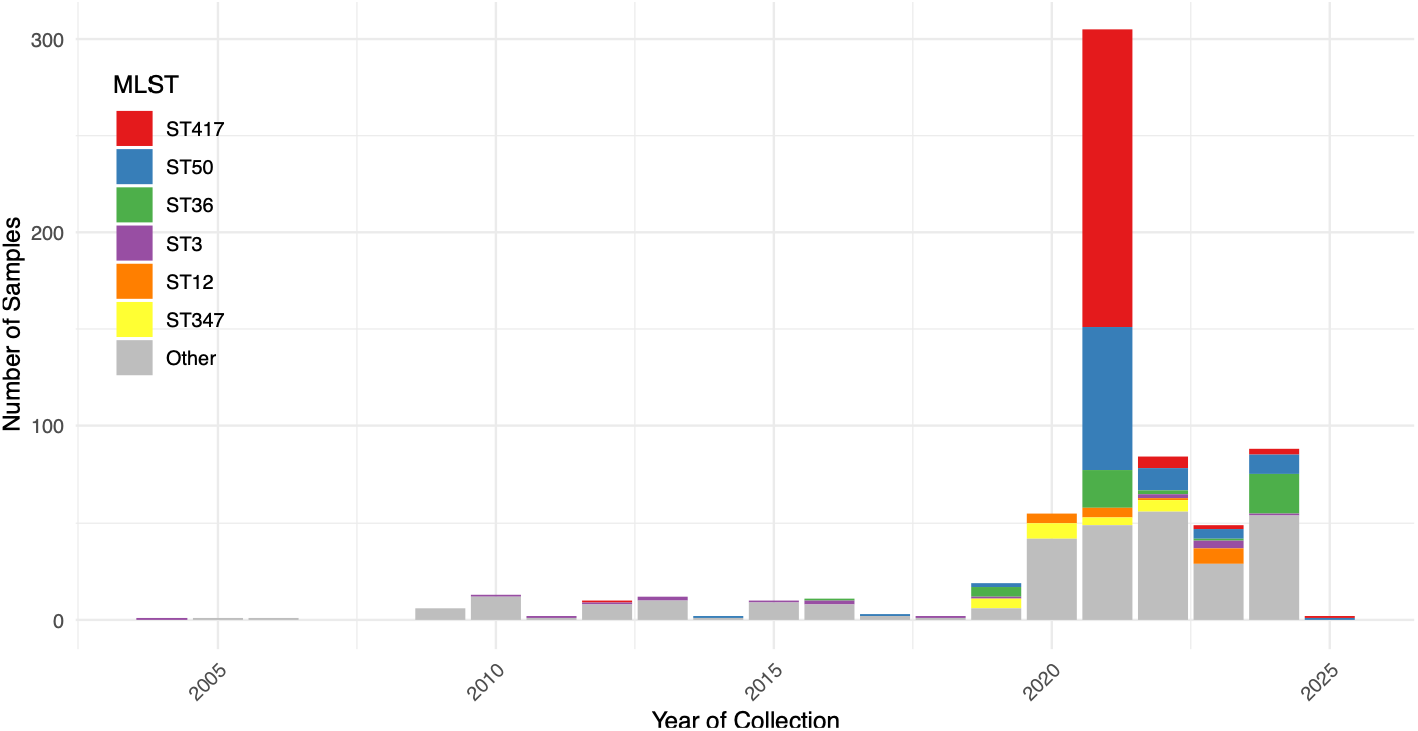
Showing counts V. parahaemolyticus genome by year of collection and coloured by MLST classification

Of the 676 *V. parahaemolyticus* isolates, 378 (56%) harboured toxin genes (toxigenic; 45 *tdh+*, 172 *trh+*, 161 *tdh+/trh+*) and 298 (44%) lacked both toxin genes. Toxigenic strains predominated among clinical cases (93.9%, 341/363) but were uncommon in environmental/food sources (10.4%, 31/299). Toxigenic isolates were distributed across 18 MLSTs. Only five sequence types (ST417, ST50, ST36, ST3, ST1140) were shared between clinical and environmental/food sources. Among these, pandemic sequence types, ST3 (n = 18) and ST36 (n = 48) were detected, while regionally dominant outbreak clones ST50 (n = 105) and ST417 (n = 167) drove the 2021 national outbreak. Additional toxigenic strains identified included ST65 (n = 5), ST817 (n = 3), and ST1140 (n = 12); ST1140 was widely detected in oysters and linked to two sporadic gastroenteritis cases. ST3 was exclusively linked to travel/imported products, with no local environmental detection. In contrast, ST36 (n = 4), ST50 (n = 4), ST417 (n = 2), and ST1140 (n = 10) were recovered from local oyster production areas, confirming environmental reservoirs. Nine other toxigenic STs (ST8, ST65, ST199, ST332, ST817, ST1107, ST1815, ST2516, ST2909) comprising 1–5 isolates each were clinical-only, and three (ST331, ST2636, STnovel) (1–3 isolates each) were environment-only. Other virulence factors including type III secretions systems (T3SS) were screened. All strains were found to carry the type 3 secretion system T3SS1, while the T3SS2 was only found in 48, 43 of which were *tdh+* carrying strains (ST3, ST8, ST1140, ST331, ST332, ST768, ST2516).

### Global genomic epidemiology and strain diversity relative to Australia/NZ

We analysed an additional 10,437 publicly available *V. parahaemolyticus* genomes (1,419 MLSTs/1,063 poppunk groups; 1951–2024) to assess the global prevalence of the key toxigenic clones (**Figure 3**). These included ST50 (n=76), ST417 (n=66), ST36 (n=801), ST3 (n=2,812), ST65 (n=13), ST817 (n=6) and ST1140 (n=10). Apart from ST3 and ST36, most toxigenic clones of interest in Australia/NZ are rarely reported elsewhere and found at low prevalences in public datasets. ST3 and ST36 dominate the genomes globally, with most originating from surveillance in USA and China. Phylogenetic analysis of the global population shows a deep, star-like branching pattern suggesting early radiation into many distinct genotypes, and geographic mapping indicates that while some have spread globally, most remain strongly tied to geographical regions. Toxin genes are found across many distinct STs, including environmental and food-derived isolates (n=869, spanning 133 STs) and clinical samples (n=5,339, spanning 177 STs). Of the 6,208 (59% of total) toxigenic genomes (carrying *tdh*+ and/or *trh*+), ST3 (*tdh*+ only) accounts for 44% and ST36 (*tdh+/trh+)* for 12% of all toxigenic genomes emphasising their global concern. 226 MLSTs each include fewer than 10 genomes. This dataset was used to place Australian isolates and lineages of concern in a global context, confirming their links to enhanced metadata and identifying localised signals.

**Figure 3.**
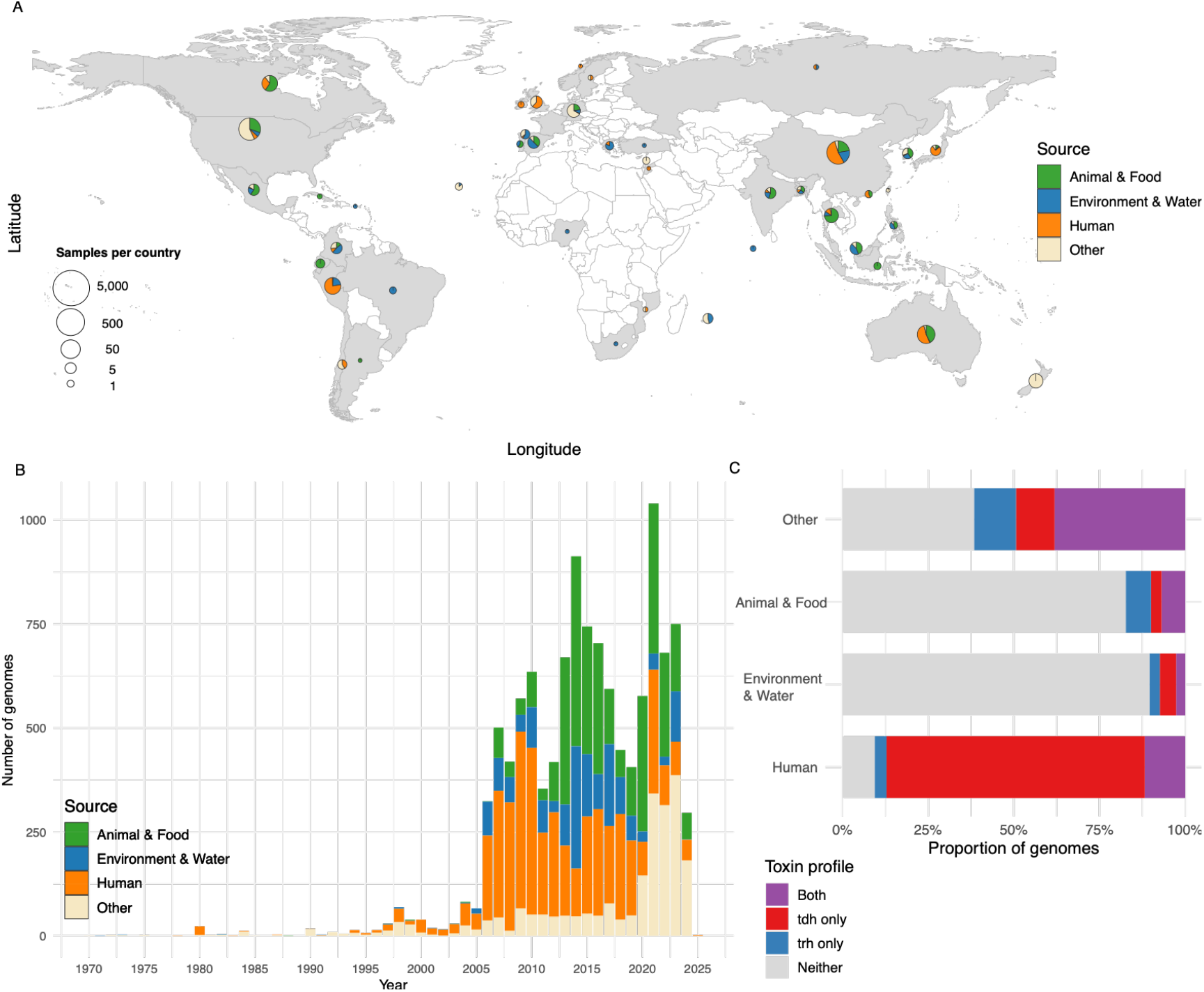
**A**. World map showing country of isolation for publicly available V. parahaemolyticus genomes coloured by source (Animal and Food (green), Environment and Water (blue), Human (orange) and Other (beige). **B**. Timeline of genomes by date of isolation coloured by source (Animal and Food (green), Environment and Water (blue), Human (orange) and Other (beige). **C**. Proportion of toxin gene positive genomes by source (No toxin (grey), trh positive (blue), tdh positive (red), both trh and tdh positive (purple).

### Emergence and spread of pandemic ST36 into Australia/NZ

Genomic analysis of the 48 Australian (from 6 states) and 396 global (7 countries) *V. parahaemolyticus* ST36 (OL4, K12) isolates indicates a single introduction from the U.S. Pacific Northwest into Australia/NZ between 2005 and 2011 (**Figure 4**). Time-scaled phylogeny shows Australian and NZ isolates forming a distinct, monophyletic clade with no mixing from other regions, supporting the emergence of a locally circulating sub-lineage in Australia/NZ. This lineage appears to have arisen from a different Pacific Northwest subgroup than those previously introduced to Spain and Peru, and is also distinct from Atlantic Northeast lineages (**Figure 5**).

**Figure 4.**
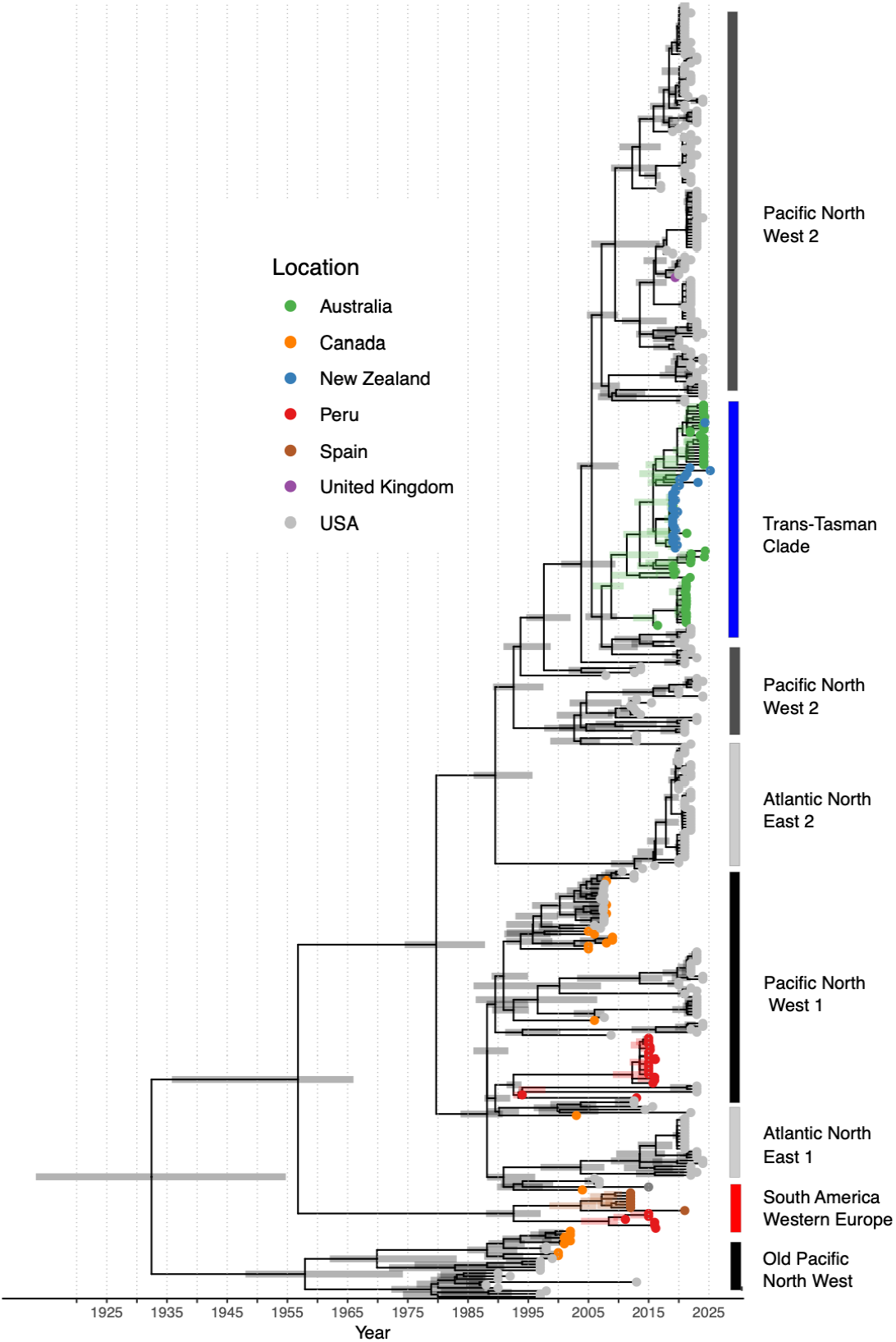
Time-scaled maximum-likelihood phylogeny of Australian V. parahaemolyticus ST36 with global context. The tree was built from 1,007 recombination-free SNPs across 506 parsimony-informative sites using IQtree2v2.1.4-beta and time-corrected nodes and branches were constructed in Treetime. Tip colours indicate country of origin (red = Peru, brown = Spain, grey = USA, orange = Canada, blue = NZ, green = Australia). Heatmap shows clade designations based on the dominant geographic origin of isolates within each clade.

**Figure 5.**
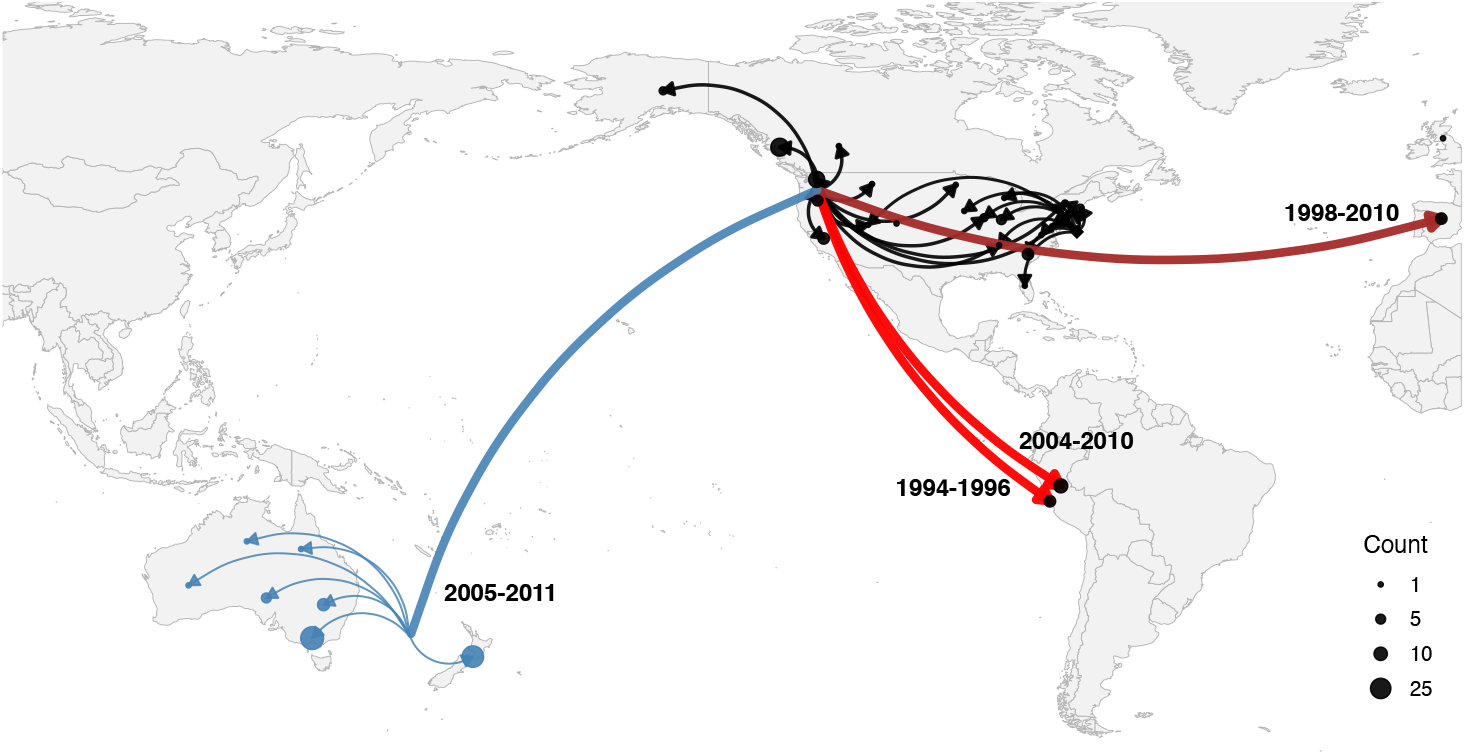
World map showing the spatial phylogenetic reconstruction of V. parahaemolyticus ST36. Blue lines indicate migration to the Trans-Tasman region, brown lines show spread to Spain, red lines to Peru, and black lines represent internal movement within the USA and Canada.

After its introduction, the ST36 lineage spread largely undetected across the Tasman Sea, causing NZ’s first well-defined domestic outbreak (May–June 2019) and two distinct Australian outbreaks (South Australia 2021; New South Wales 2024)^11,12^. Cluster analysis differentiates the 2019 NZ outbreak, the 2021 outbreak (VIC/SA), and the 2024 outbreak (VIC/NSW) as separate but closely related subgroups (≤5 SNPs) within the broader Trans-Tasman clade. This suggests that a single-digit SNP threshold combined with epidemiological metadata can serve as a pragmatic baseline for reliable outbreak detection.

### Toxigenic ST50 and ST417 show evidence of regional persistence

Time-scaled phylogeny of *V. parahaemolyticus* ST50 (O6:K18) including 105 Australian isolates from six states and 121 global isolates (82 from NZ) shows a large monophyletic, Trans-Tasman clade representing most ST50 cases in the region (**Figure 6A, 6C**). Within this clade there are two major groupings that broadly align with the outbreaks reported in Australia and NZ, although there is some intermixing between countries with isolates present across the groups from non-outbreak linked cases from other time periods outside the outbreak windows. Within this clade, the most recent common ancestor (MRCA) of the NZ outbreak and surveillance isolates dates to late 2019, while the Australian subclade traces back to 2018 (**Figure 6C**). The close genetic relatedness between Australian and NZ isolates suggests clonal expansion from a single regional source.

**Figure 6.**
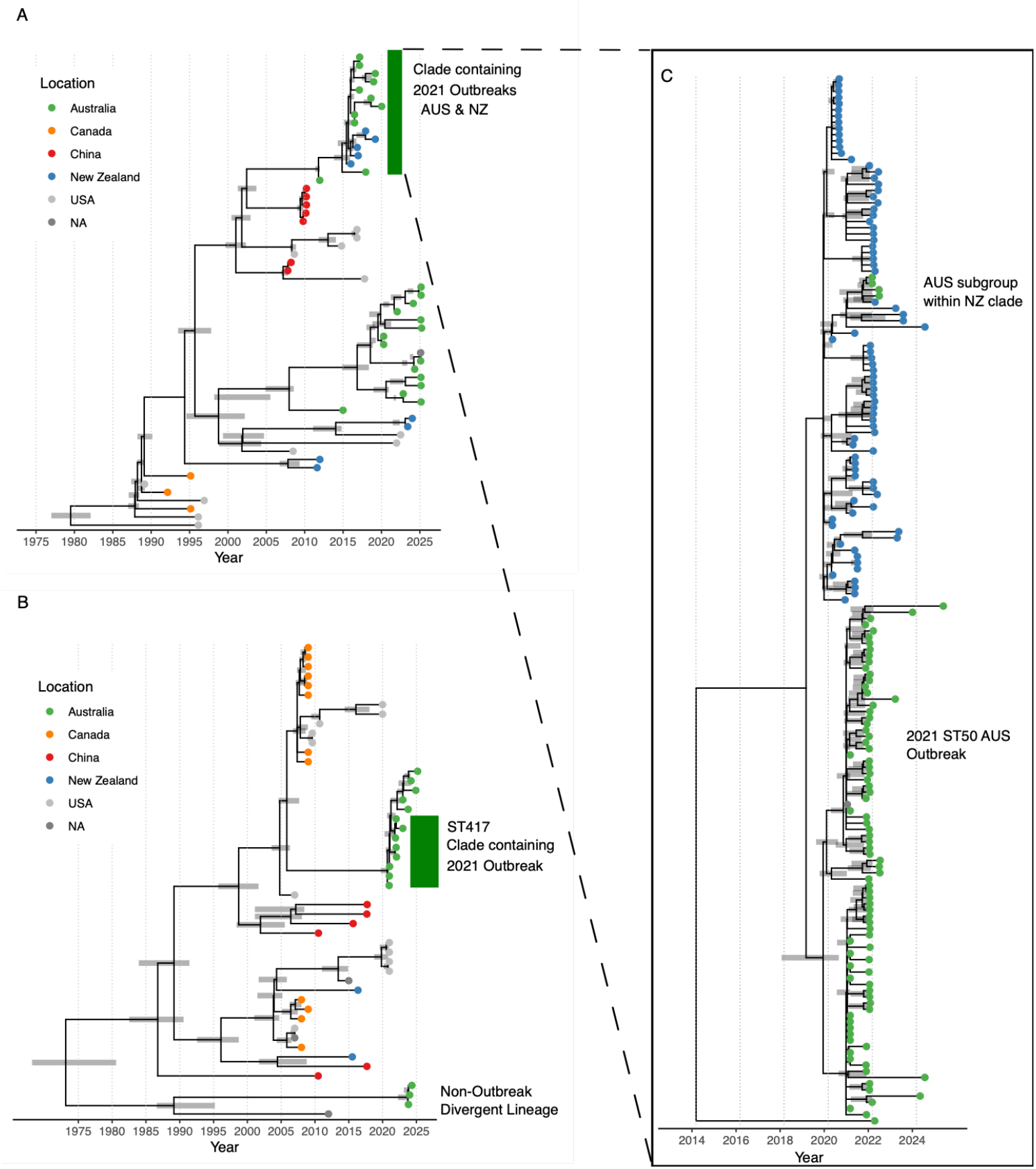
**A**. Time-scaled maximum likelihood phylogeny of ST50 V. parahaemolyticus with global isolates. The tree was built from 959 recombination-free SNPs across 484 informative sites using IQtree2 and time-corrected nodes and branches were constructed in Treetime **B**. Time-scaled maximum likelihood phylogeny of ST417 V. parahaemolyticus with global isolates. The tree was built from 440 recombination-free SNPs across 219 informative sites using IQtree2 and time-corrected nodes and branches were constructed in Treetime. **C**. Time-scaled maximum likelihood phylogeny of ST50 the Trans-Tasman region Clade. The tree was built from 129 recombination free SNPs across 43 informative sites using IQtree2 and time-corrected nodes and branches were constructed in Treetime. Tip colours indicate country of origin (red = China, grey = USA, orange = Canada, blue = New Zealand, green = Australia, dark grey = unknown).

Comparison with the global context genomes shows that ST50 is genetically diverse and has likely persisted regionally for a period before the recent outbreaks (**Figure 6A**). Several Australian and NZ isolates fall outside the Trans-Tasman clade, forming highly divergent lineages (80–5,000 SNPs across the 91% core genome) interspersed with isolates from other regions (e.g., USA, China). These outlier groups suggest long-term local diversification and undetected movement within the Trans-Tasman region.

We applied the same approach to the ST417 isolates (**Figure 6B**). Genome comparisons demonstrated the outbreak clade consists of an Australian monophyletic cluster (O1:KUT2) with no close matches among available global genomes (China, Canada, USA, Thailand, NZ), and that remains ∼500–1,500 SNPs away from the rest of the ST417 isolates. After the outbreak, closely related sequences have been identified in sporadic cases; however, none have been associated with a subsequent large-scale outbreak, indicating a persistence of these genotypes within the environment. Three ST417 genomes with a different serotype combination (O1:K25) form a highly divergent secondary cluster to the outbreak clade. In comparison to ST50 only two genomes of ST417 were available for NZ and represent sporadic non-outbreak associated cases.

## Discussion

The number of reported *V. parahaemolyticus* outbreaks in Australia and NZ has increased over the past decade^10,27^. While environmental factors and improved detection and reporting have likely contributed to this trend, introduction and regional spread of specific genetic lineages have also played a key role^28^. We demonstrate how the adoption of WGS has been critical for understanding the global movement and diversification of pathogenic *V. parahaemolyticus*. Our study demonstrates ST36 was likely introduced from the U.S. Pacific Northwest into the Trans-Tasman region^29^, where it has since diversified locally and driven multiple outbreaks. Similarly, emerging STs, ST50 and ST417 appear to have expanded regionally from an endemic diverse population, highlighting the importance of genomic surveillance for detecting and tracking clones of concern.

ST36 has recently emerged in Australia and NZ as a significant cause of seafood-associated outbreaks of human gastroeneritis^13^. Historical records show very few outbreaks in Australia and NZ, with no prior detection of ST36 in the region, but since 2010 ST36 has been increasingly identified in both clinical and environmental samples. Genomic analysis suggests a single introduction from the U.S. Pacific Northwest between 2005 and 2011, followed by local circulation, diversification, and adaptation that contributed to multiple foodborne outbreaks in Australia and NZ (**Figures 4, 5**). This pattern mirrors ST36’s history of repeated long-range spread and regional establishment^14,15,29^. The Pacific Northwest is recognised as the origin of ST36, with strains dispersing to the U.S. Northeast in the mid-1990s, where they diversified, became resident, and caused repeated oyster-associated outbreaks^14,15,30^. Multiple introductions into Peru (late 1990s and 2011–2016)^29^ and Spain (2012)^31^ also seeded local populations in regional waters triggering outbreaks. The estimated arrival of ST36 in the Trans-Tasman region predates the first recognised outbreaks, suggesting undetected environmental persistence for several years before emerging as a public health concern when suitable environmental conditions enabled outbreaks to occur.

These findings fit within a broader global pattern of *V. parahaemolyticus* lineages crossing oceans and adapting to new environments. The earlier ST3 O3:K6 pandemic clone demonstrated the ability of *V. parahaemolyticus* to move internationally and establish stable reservoirs^16,32^. ST36 shows a similar trajectory but with multiple independent transoceanic introductions from the Pacific Northwest into the Atlantic, South America, Europe, and now the Trans-Tasman region^15,28,33,33^. Its capacity to colonise new coastal ecosystems, diversify locally, and fuel recurrent seafood-borne outbreaks underscores its pandemic potential. Understanding the timing and pathways of these movements, likely linked to global seafood trade^34^, transport via ballast water, bird migrations^35^, seasonality^36^ and/or climate-driven changes in marine environments^37,38^ is essential. Expanding environmental and genomic surveillance will help track and determine the time of future migrations, characterise local adaptation, and guide early detection and outbreak response.

ST50 and ST417 reflect two distinct lineages, with similar evolutionary patterns, established in the Trans-Tasman region. ST50 shows a clear pattern of local expansion from a single recent source, driving the major NZ and Australian outbreaks between 2018–2021^8,10,13^ (**Figure 6A, C**). This expansion occurs against a much older, genetically diverse background likely established decades earlier, suggesting long-term persistence of this ST in regional marine environments, with periodic emergence of human infections and outbreaks. In contrast, ST417 displays a strongly Australian-confined signal, with only sporadic cases in NZ (**Figure 6B**); its large outbreak cluster sits alongside highly diverged local outliers, and its broad common ancestor range plus genomic distance from overseas isolates indicate a long-standing, diverse Australian population that only recently produced an outbreak-associated clone.

The patterns described in this study indicate ST50, ST417, and ST36 may be STs that are more adapted to cooler marine waters. Experimental work shows ST36 can grow in oysters at low temperatures previously considered unsuitable for *V. parahaemolyticus*^2,39,40^. All three of these STs have emerged as key strains of concern in the past decade, while other toxigenic strains such as ST3 have not become established in Australian waters, raising important questions about why these clones, and not others, have successfully persisted and driven outbreaks in the region.

This study provides the first genomic baseline for *V. parahaemolyticus* in Australia, supporting interpretation of cases under the national vibriosis notification framework introduced in January 2025. The results emphasise the importance of national coordination, traceability and a flexible outbreak definition that reflects interstate seafood trade and the polyclonal nature of outbreaks and transmission, where MLST/serotyping and toxinotyping alone lacks resolution within dominant persistent STs such as ST36, ST50 and ST417.

In conclusion, understanding the regional endemicity of outbreak-causing and high-risk toxigenic *V. parahaemolyticus* strains is key to assessing introduction risk and local transmission potential. Differentiating between infections caused by locally contaminated seafood and those linked to imported products or travel informs both risk assessment and control strategies by government agencies and seafood industry. Targeted local environmental screening helps identify strains circulating within a region and those most likely to cause disease, while accurate travel history and source attribution reduce the risk of mistakenly linking imported cases to local production. Combining enhanced metadata with WGS provides a robust framework for source tracking and outbreak response, which will be essential for managing this foodborne disease in an increasingly interconnected and dynamic One Health system, where environmental reservoirs, food production, and human movement collectively shape the emergence, spread, and persistence of clinically significant *V. parahaemolyticus* lineages.

## Data Availability

All data produced in the present work are contained in the manuscript

https://github.com/JA-Lacey/SafeFish-Vpara

## Funding Source

This work was funded by The Commonwealth Government of Australia through Safefish 2021-2025 (Fisheries Research and Development Corporation Project 2021-018).

## Acknowledgments

We thank the laboratory staff at UTAS IMAS, South Australian Research and Development Institute (SARDI), EMAI, SA Pathology, PathWest Laboratory Medicine WA, Institute of Clinical Pathology and Medical Research (ICPMR) NSW Health Pathology, Public Health Microbiology Public and Environmental Health Reference Laboratories (Queensland), Tasmania Department of Health, and Microbiological Diagnostic Unit Public Health Laboratory (MDU-PHL) for their role in sample testing, analysis and reporting.

## Notes

### Competing Interest Statement

The authors have declared no competing interest.

### Author Declarations

Human Research Ethics Committee of The University of Tasmania Waived ethical approval for this work.

